# Beyond vaccination: A Cross-Sectional Study of the importance of Behavioral and Native Factors on COVID-19 Infection and Severity

**DOI:** 10.1101/2022.01.23.22269214

**Authors:** Hani Amir Aouissi, Mostefa Ababsa, Carlos M. Leveau, Alexandru-Ionut Petrisor, Artur Słomka, Mohamed Seif Allah Kechebar, Jun Yasuhara, Loïc Epelboin, Norio Ohmagari

## Abstract

The COVID-19 pandemic has a major impact on a global scale. Understanding the innate and lifestyle-related factors influencing the rate and severity of COVID-19 is important for making evidence-based recommendations. This cross-sectional study aimed at establishing a potential relationship between human characteristics and vulnerability/resistance to SARS-CoV-2. We hypothesize that the impact of virus is not the same due to cultural and ethnic differences. A cross-sectional study was performed using an online questionnaire. The methodology included a development of a multi-language survey, expert evaluation and data analysis. Data was collected using a 13-item pre-tested questionnaire based on a literature review. Data was statistically analyzed using the logistic regression. For a total of 1125 respondents, 332 (29.5%) were COVID-19 positive, among them 130 (11.5%) required home-based treatment, and 14 (1.2%) intensive care. The significant factors included age, physical activity and health status all found to have a significant influence on the infection (p < 0.05). The severity of infection was associated with preventive measures and tobacco (p < 0.05). This suggests the importance of behavioral factors compared to innate ones. Apparently, the individual behavior is mainly responsible for the spread of the virus. Adopting a healthy lifestyle and scrupulously observing preventive measures including vaccination would greatly limit the probability of infection and prevent the development of severe COVID-19.

## 1. Introduction

### 1.1 Background

According to the WHO (World Health Organization), several coronaviruses (CoVs) have been reported since 2002. The coronavirus responsible for the 2019 pandemic was named SARS-CoV-2 by ICTV (International Committee on Taxonomy of Viruses) due to its similarity to SARS viruses [1].

In the end of 2019, a novel beta coronavirus was identified in China’s Hubei province, at Wuhan [2]. By that time, the virus has spread and disrupted all aspects of humans worldwide. The symptoms of severe acute respiratory syndrome coronavirus 2 are similar to those of previously known coronavirus infections including fever, dry cough, and fatigue; however, the SARS-CoV-2 has a higher spreading nature [3,4]. It has largely surpassed MERS and SARS in terms of both spatial range of epidemic areas and number of infections. The global health crisis experienced has posed a significant threat to public health [5].

According to the WHO’s weekly operational update on COVID-19 of 13 December 2021, there are almost 273 million confirmed cases and 5,34 million related deaths worldwide [6]. The consequences were serious, especially at the start of epidemic, given the lack of information and means to fight this virus [7]. Across the world, a significant number of proactive measures have been adopted, including distance education, banning international traveling, and the campaign encouraging everyone to “stay at home” [8]. Given the unknown and unexpected aspect of this epidemic in modern times, there is inescapably a lack of research on its psychosocial effects [9].

Several remedies have been proposed over time to try to eradicate this virus, or limit its spread. The most famous examples are Chloroquine (CQ) and Hydroxychloroquine (HCQ) with or without Azithromycin, still used today as the last alternative in some countries [10–12]. Other notable examples of proposed remedies include Remdesivir [13,14], Favipiravir [15], Dexamethasone [16], Artemesnin [17] and Ivermectin [18]. Actually, as many clinical trials are still repurposing existing drugs [19]. Since the publication of the genetic sequence of SARS-CoV-2, the whole world has actively started trying to produce vaccines [20,21]. At present, out of a total of 8,65 billion administrated doses, more than 3,63 billion people are already fully vaccinated, representing more than 46.6% of the world population [22]. Each of the vaccines administered worldwide, actually has been subject to specific studies: Johnson & Johnson [23,24], Pfizer/BioNTech [25,26], Moderna [27,28], AstraZeneca [29,30], and Sputnik V [31,32]. China has around fifteen vaccines. In addition to being the first developed, they are the most numerous ones [33,34]. The most famous are CanSino [35], Sinopharm [36], and finally Sinovac [37]. The latter, in particular, was subject to very complete studies; the first one demonstrated that this vaccine effectively prevented the virus, including severe disease and death [38]. The second one compared the effectiveness of this vaccine to Alpha and Delta variants, and demonstrated only a meager difference in vaccination effectiveness with the Delta variant compared with the Alpha variant after the receipt of two vaccine doses [39]. The third one showed low efficacy against the omicron variant [40]. The possibility of mixing several vaccines is also being considered [41]. Currently, the possibility of a fourth dose is considered in some countries especially since the appearance of the Omicron variant [42].

Understanding the innate and lifestyle-related factors on the incidence and severity of COVID-19 is critical for making appropriate recommendations to prevent the transmission of COVID-19 as well as the development of severe COVID-19. However, the detailed characteristics influencing the rate and severity of COVID-19 is not fully understood. Previously, the published studies were mainly interested in the influence of COVID-19 on the changes of lifestyle and not the opposite [43,44]. So far, the COVID-19 questionnaires related much more to the perception of people concerning this epidemic [45]. Since the epidemic had occurred, several international questionnaires were created. Their purpose was to answer specific question, e.g., about the knowledge and attitude of residents in the prevention and control of COVID-19 [2], or the preventive practices against COVID-19 pandemic in the general population [46]. They were about the mental health impact on people with and without depressive, anxiety, or obsessive-compulsive disorders [47], eating habits, activity and sleep behavior [48], etc. Others focused on particular countries like China [49], USA [50], and France [51]. Herein, we conducted a cross-sectional analysis to evaluate the influence of innate and lifestyle characteristics on COVID-19 by using an online questionnaire based on the literature review as follows.

### 1.2. Literature review on the main risk factors

A comprehensive literature review using databases and search engines, such as the Web of Science, Google Scholar, PubMed, and Scopus, was carried out to acquire a global overview of the relationships between the different risk factors and COVID-19. The following keywords: “coronavirus”, “COVID-19”, “questionnaire”, “survey”, “tobacco”, “alcohol”, “lifestyle”, “behavior”, “immunity”, “ethnic origin”, “continent”, “country”, “ epidemic”, “blood group”, “sports activity”, “age”, “educational attainment” and “preventive measures” were included in our search. The search yielded more than 201 related articles. Furthermore, after screening the titles, abstracts, and full contents, approximately 170 articles were found relevant. 64 were used in the literature review.

The literature review allowed for identifying the risk factors described in the introduction. The conclusion of several studies has amply demonstrated that changes in individual factors would directly (or indirectly) affect the spread of virus. Through literature review and expert opinion, some factors were found to be determinant, e.g. age, social distancing, air temperature, ventilation/airflow, humidity, population density, and community consciousness. These factors were interdependently found to have a strong impact on the SARS-CoV-2 epidemic characteristics [52,53].

#### 1.2.1 Tobacco

According to the WHO, tobacco is one of the main causes of premature death and morbidity. It is known that smoking and consuming smokeless tobacco (SLT) products significantly increases the risk of NCDs (non-communicable diseases). According to Islam and Walton [54], only few studies had investigated the relationship between tobacco consumption and COVID-19 [55]. Some of them have showed that the hospitalization rate of smokers is higher than that of non-smokers [56,57]. Berlin et al. [58] encouraged stopping smoking and repeated gestures that facilitate contamination, and called for the awareness of health authorities. Mistry et al. [59], in their study, found an increased risk of mortality specifically for people over 65, especially those who live in urban areas, because they are exposed to pollution. Yingst et al. [55] stated that that the increased stress can be detrimental, and it is imperative to create innovative methods to support users interested in quitting during this particularly difficult period.

#### 1.2.2 Alcohol

Alcohol consumption increases the risk of community-acquired infections [60]. Nonetheless, news and media showed that the false propaganda of racketeers misled the public into believing that, since alcohol can remove viruses, its consumption can destroy the coronaviruses after entering the body, supposedly because ethanol is commonly used for hand sanitizers [61], but inescapably this is nothing more than a myth. Some studies demonstrated that alcohol poisoning, which is a consequence of the increase in alcohol consumption, increased significantly especially with the appearance of SARS-CoV-2 epidemic [62,63]. In most communities, alcohol poisoning is responsible for several health issues although it can easily be avoided [64]. The availability of methanol and ethanol in recent years led to an augmentation of morbidity and mortality due to alcohol poisoning in some low-income countries, specifically the Muslim ones [65]. Lassen et al. [66] demonstrated that weekly alcohol consumption is associated with the progression to ARDS during hospitalization with COVID-19, but there is a huge lack of studies dealing with alcohol consumption and infection by SARS-CoV-2.

#### 1.2.3 Physical activity

Regular sports activity is associated with a decrease of 6.0% to 9.0% of the risk of influenza-associated mortality [67]. It was demonstrated previously, through a study on mice that exercise (moderately) in the first days after an influenza virus infection, that the mortality was reduced [68]. However, it remains unclear how the physical exercise affects infections. Endurance exercise that lasts less than 1 hour stimulates NK cell cytotoxicity [69]. Moreover, mild physical activity may boost the immune system, whilst exhausting exercise may weaken it [70]. In general, an unhealthy lifestyle could have negative consequences, whether dealing with the virus now or in the post-COVID-19 period, specifically in sedentary people. The consequences that a sedentary situation can cause and which we can observe over time are, inexorably, cardiovascular diseases, like obesity, diabetes, hypertension, and metabolic syndrome, because a decrease in activity is logically associated with a reduction in insulin sensitivity according to Narici et al. [71]. The same authors claimed that inactivity during confinement could result in a reduction of cardiac volume, oxygen absorption capacity, and VO2max (maximum oxygen consumption). A reduction in VO2max and the absorption capacity of oxygen (O2) is often associated with a high mortality rate. In addition, this decrease affects blood circulation and the oxidative function of muscles. In these circumstances, sports activity can be defined as a key strategy to fight against unhealthy lifestyle during the pandemic or even after [72] since it contributes to maintaining an optimal state of health (mental/physical). The WHO proposed many general recommendations for sports activity to combat the confinement situation. It consists of performing 150 min or at least 75 min/week of physical activity (moderate or vigorous). It is also recommended to combine activities of different intensities [73].

#### 1.2.4 Blood group

The majority of studies reported that group A had an increased frequency amongst SARS-CoV-2 infection risk, and the opposite for group O, which is associated with a decreased frequency and risk of infection [3,74]. However, recent studies have shown neither an association between blood group and COVID-19 infection susceptibility nor between blood group O and a decreased risk of infection [75]. The role of blood group in the infectivity of SARS-CoV-2 and COVID-19 severity requires further studies. The role of ABO type is likely secondary and non-modifiable [76] and, apart from blood groups, other population-dependent antigens distribution may be of importance in the fairly heterogeneous spread of the virus in all the geographic areas [77].

#### 1.2.5 Ethnicity

Ethnicity is a complex concept, which can include several multiple dimensions, as it is socially constructed and may be associated with biological attributes, such as skin color [78]. Black, Asian, and minority ethnic communities in the UK made up 36.0% of the critically ill patients with COVID-19 requiring intensive care [79]. This phenomenon is not specific to UK; it turns out that this is usually the case in several countries. There is considerable evidence that the incidence of clinically important diseases is higher in many minority ethnic groups despite the fact that impacts of race/ethnicity are likely to vary in different countries, as underlined by some studies [80,81].

Some meta-analyses were published recently. According to Sze et al. [82], there is a clear confirmation that patients of Asian, Black, and Hispanic ethnicities have an increased probability of being infected with coronavirus, compared to white ethnics. In addition, there is a possibly higher risk of intensive care admission and death in Asians. Their findings should be used by institutions and decision-makers to minimize the exposure to COVID-19 in ethnic minorities. This can be achieved through some measures such as helping them access health care resources quickly, observing ethical standards and eliminating as much as possible all forms of racism, social determinism, and inequalities. For Vist et al. [83], even if there is an increased risk of infection and hospital admission due to SARS-CoV-2 for several immigrants and minority ethnic groups, this is especially associated with a low socioeconomic status, resulting into higher admission to hospital and death rates compared to groups with a high socioeconomic status. The fact that may explain the tragic situation in some countries compared to others with almost similar ethnicity [84].

#### 1.2.6 Gender

Generally, data around the world indicate higher COVID-19 case fatality rates among men than women [85]. Gender-based behavioral and socio-cultural differences may contribute to the sex differences reported in the severity of COVID-19 disease. For example, men wash their hands lesser, especially with soap or other products after entering a restroom, and are more likely to smoke [86].

A large-scale data of two meta-analysis studies demonstrated that despite the fact that there are no significant differences in the proportion of infected individuals with the virus, men are much more likely to develop serious illnesses and die compared to women [87,88] except for some countries like India [89]. According to Bhopal and Bhopal [90], hypotheses based on risk factors that are known to change with both sex and age seem to be the most probable explanations for the observed differences. The differences are due to lifestyle (e.g., alcohol, smoking), occupation, use of medications, or medical co-morbidities. These descriptions reflect cultural and social characteristics associated with gender rather than the biology of sex.

#### 1.2.7 Age

Since the beginning of the epidemic, age has been defined as the keystone in COVID-19 infected people [91]. The first data coming from Italy also attested that mortality was highly significant in septuagenarian patients and even higher in octogenarians [92]. A meta-analysis has suggested an essential influence of age on mortality of infected patients with a relevant threshold on age>50 and more especially>60 [93]. This epidemic reminds us daily that older individuals, specifically those with rheumatic diseases [94], are extremely vulnerable to infectious diseases due to co-morbidities linked with age and a decrease in immunological competence or “immunosenescence” [95]. This process, particularly the increased production of inflammatory cytokines resulting from inflammation, is partly responsible for determining the prognosis of COVID-19 in elderly individuals. [96].

The peculiarities of the immune system of older individuals may contribute to both the deficiency of effector mechanisms essential for viral pathogens fighting and an exacerbated inflammatory response, which can accelerate and intensify lung tissue damage [97]. Nonetheless, several authors agree on the fact that one should in no way consider COVID-19 as a “disease of the elderly”, but the pandemic should be considered as a temporary setback in a long-term decline in senescent mortality [98].

#### 1.2.8 Resistance (immunity)

The genetic background of patients and presence of concomitant pathological conditions were also examined, provided that adaptive immunity to closely related viruses or other microorganisms can reduce susceptibility or increase the severity of disease [99]. In line with some studies, seemingly most severely affected people have misguided antibodies (autoantibodies) that attack the immune system rather than the virus that causes the disease. A small percentage of the population who develop severe COVID-19 is characterized by a specific genetic mutation that affects immunity. Consequently, many individuals lack effective immune responses, which depend on type I interferon [100,101].

At the same time, it seems that a part of population is more resistant to COVID-19, for example, according to the study by Le Bert et al. [102], some have developed immune cells called T lymphocytes, which are directed against the new coronavirus. This immune response would be “lasting”, unlike antibodies, which would disappear quickly. Above all, this response would be present in more than 50.0% of healthy people who have never been infected with the coronavirus. It also demonstrated that different efficacy of both innate and immune responses, according to age and co-morbid conditions, can be a confounding factor [103]. Grasping the combinational and individual role of hosts, environmental factors, and viral ones in COVID-19 infection provides a better knowledge of the eventual high-risk groups of people concerning SARS-CoV-2 specifically in terms of severity and susceptibility [104].

#### 1.2.9 Educational attainment

It has been demonstrated that educational level has an effect on the incidence of some diseases (specifically non-communicable). Low levels of education are related to high prevalence and incidence of cerebrovascular and cardiovascular diseases, cancer, diabetes, hypertension, and chronic respiratory diseases [105]. Generally, a bad socioeconomic status may be linked to low educational level, thus increasing the risk of previously mentioned diseases [106], and of COVID-19 because of low levels of immune cells and high levels of Cytokines in body fluids [107]. It should be remembered that the access to education is not the same elsewhere; when examining urban versus rural outcomes in COVID-19, the disparities are markedly evident when considering county-level data [108]. Concerning the relationship between COVID-19 and educational level, it is acknowledged that education and engagement in health behaviors are positively associated [109], and low education is associated with unhealthy behaviors [110]. However, several external factors may be examined and additional measures will have to be studied each time to test for this relationship [111]. Some studies have also suggested that a standardized measurement should be used in future studies [112].

#### 1.2.10 The continent of residence

As a pandemic, COVID-19 had hit almost every country on earth. Different approaches to emergency management were adopted in each country [113] as in East Asia [114]. Some studies analyzed the differences and similarities in how the inhabitants of different countries responded to government policies enacted during the spread of the COVID-19 pandemic [115] by measuring for example confirmed cases and deaths, the stringency index. The results varied according to each country [116]. If possible, a more government-related response would be worth investigating. Nevertheless, it has been demonstrated that the lag time in responding to government policy and response variability in each country are significantly correlated with the numbers of infections and deaths [117].

#### 1.2.11 Observance of protective measures

According to the WHO [6], there are many health precautions that individuals can take to avoid COVID-19 infection including frequent hand-washing, wearing facial coverings, maintaining a 2 m distance between themselves and others, staying away from crowds, coughing/sneezing into the elbow (or mask), cleaning/disinfecting surfaces regularly, and examination by a doctor if necessary as recommended by the CDC [118]. In addition, we included vaccination among the strict preventive measures given that at the time when the questionnaire was created, the vaccination process was barely starting and its contribution almost negligible. However, we still have considered it as an additional measure, and discussed in more detail in the introduction.

### 1.3. Aims and importance of research

Given the ambiguity of answers from previous studies that used questionnaires, the present research was designed to correct some past limitations. The aim of this study was to investigate the effects of factors beyond human control (e.g., blood group, ethnicity) on COVID-19 potential infection and severity, and of lifestyle or behavioral characteristics (e.g., tobacco use, sports activity, and alcohol consumption). Based on the fact that the number of infections varies greatly by country and continent (even if this may also be due to significant variations in the level of underreporting of cases between countries), we hypothesized that the innate factors should be the most significant in deciding on the possible infection of individuals. We examined this assumption using questionnaires for quantification.

## 2. Materials and Methods

### 2.1 Overview

The present study is based on a cross-sectional design, using an online questionnaire. The main function of questionnaires was to give the survey a greater extension. It was also used as a means to verify statistically the extent of generalization that can be reached by the existent information and preliminary assumptions [119].

To accomplish the aim, we designed an anonymous online survey with a sufficient number of items; its qualities include conciseness, scientific structure, pre-testing and global practicability. Considering the lack of similar works, the questionnaire addressed some basic information and all major lifestyle aspects. To ensure optimal compliance with preventive measures against the spread of virus, we have deliberately chosen to share the survey exclusively online, to limit contact as much as possible. A standardized methodology including steps such as literature review, expert review, ethical approval and pre-testing were undertaken to develop and validate the questionnaire [120,121].

### 2.2 Development of the questionnaire

We drafted our questionnaire in four languages: Arabic, English, Spanish and French, which are in the top 6 most spoken worldwide, along with Chinese and Hindi [122]. The data was collected via Google Forms, through a self-report questionnaire. Additionally, the questionnaire consisted of three main parts. The first one was related to the socio-demographic characteristics. The second part consisted of items related to behavior, and the last part included the infection status (yes or no) and its severity. We made the questionnaire as easy as possible with a minimum of items, so that it could be understood by almost the entire world population. We have particularly made sure that it is very fast to answer so as not to be discouraging, while deliberately excluding criteria that are too subjective or difficult to measure (e.g., hours of sleep, quality of nutrition, and stress rate). To highlight the importance of research and great credibility, the tool was presented to a committee of national and international experts who helped to improve it by deleting or rewording some items. Their intervention made possible to ensure its suitability and validity before applying it to the participants. Conventionally, the ethical approval and consent were obtained from the CRSTRA and all participants. The questionnaire was shared via social media including Instagram, Facebook, LinkedIn, WhatsApp and volunteer participants were requested to fill in the online form. However, we required that the respondents should be at least 18 years old to answer, and asked for a confirmation for each respondent.

### 2.3. Statistical analysis

Statistical analyses were performed using SAS® (version 9.4). We coded the contamination factors on a numerical scale due to their nature (nominal or ordinal variable). For this reason, the responses were coded as following:

A progressive scale describing the direction of variation concerning the factors of an ordinal nature:

-Educational attainment: 0-3 (0=Not precise, 1= No study or primary, 2= Middle/secondary, 3= University/post-university) - Age: 1-4 (1=18-30, 2= 31-45, 3= 46-59, 4= 60+) - Sports activity: 1-3 (1= Little or no activity, 2= Moderate, 3= Very active) - Health status: 1-3 (1=Resistant, 2= Moderately sensitive, 3= Very sensitive). - Tobacco use: 1-3 (1=No, 2= Occasionally, 3= Frequently) - Alcohol consumption: 1-3 (1=No, 2= Occasionally, 3= Frequently) - Protective measures against COVID-19: 1-3 (1=Not at all, 2= Medium application, 3= Strict application) - Severity: 0-3 (0=Healthy, 1= Low, 2= Treatment, 3= Intensive care).

A two-choice scale for the question: “Have you been affected by COVID-19?” (No=0, Yes=1).

A random scale for factors of a nominal nature: - Residence (1=Africa, 2= Europe, 3= North America, 4= South America, 5= Asia, 6= Oceania) - Ethnicity (0= Not precise, 1= Other, 2=African / Afro-American, 3= Caucasian, 4= Arabic, 5= Asian, 6= Latino) - Gender (0=Not precise, 1= Male, 2= Female, 3= Other).

We noticed that the estimations of odds ratios were affected by very small sample size for specific levels of the variables. In order to increase the validity of statistical analyses, data for some levels were removed, by changing them to missing observations: Gender – 3 “Other” values, Continent – 2 “Oceania” values, Health – 1 “0=Healthy” value, Continent – 10 “4=South America” values, and Ethnicity – 11 “6=Latino” values. However, we did not eliminate any blood groups, despite of the reduced sample size influencing in one case the estimation of odds ratios, because it make no sense from a health standpoint.

Two statistical analyses were run:

- The dependence of having been affected before or not by COVID-19 on the potential risk factors investigated was analyzed using logistic regression, a statistical method modeling a binary event as probability of its occurrence.
- The dependence of the severity of infection on potential risk factors investigated was also analyzed using logistic regression, after eliminating healthy subjects from the analysis and turning the “Severity” variable into a binary one, which indicates if the patient needs treatment or intensive care.

Provided that the difference in the odds ratios is not the same for the different levels of the predictors considered in this study, absolutely all independent variables were treated as categorical in all analyses.

Model selection was performed with the intention of finding a model including only predictors with a significant impact on the response when acting together. The model selection was performed using the approach proposed by Collett [123], consisting of: (1) performing univariate regressions with each predictor separately, (2) building a first multivariable model with all predictors if their p-values in (1) were below 0.2; (3) performing a backward elimination with the model obtained in (2), by eliminating the variable with the least influence on the response (indicated by the highest p value), until all variables in the model had a statistically significant influence on the response, and (4) attempting to re-include variables not retained at the end of (3), in order to check if the model quality can be improved.

For all statistical analysis, the level of significance was 0.05. However, since the sample size was not very large, the presentation of results also includes results significant at the 0.1 level. In this case, additional data would probably have led to significant results.

## 3. Results

### 3.1. Descriptive statistics

For a total of 1125 responses, 753 responses (67.0%) were made in French, 225 (20.0%) in Arabic, 135 (12.0%) in English, and finally 12 (1.0%) in Spanish.

**Figure 1.**
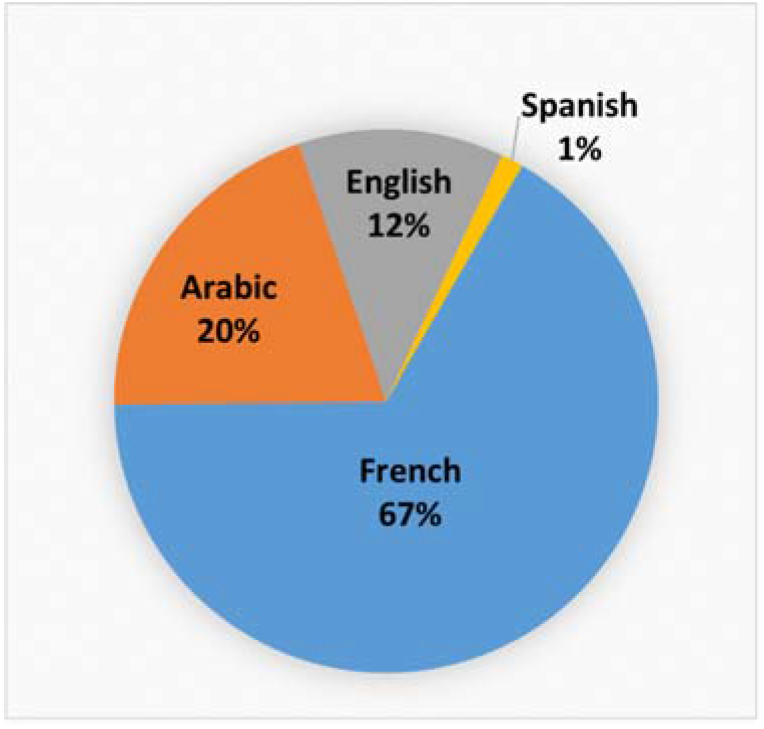
Representation of received anwsers

For 1125 respondents, 332 were COVID-19 positive, 130 required home-based treatment, and 14 required an intensive care.

**Table 1.**
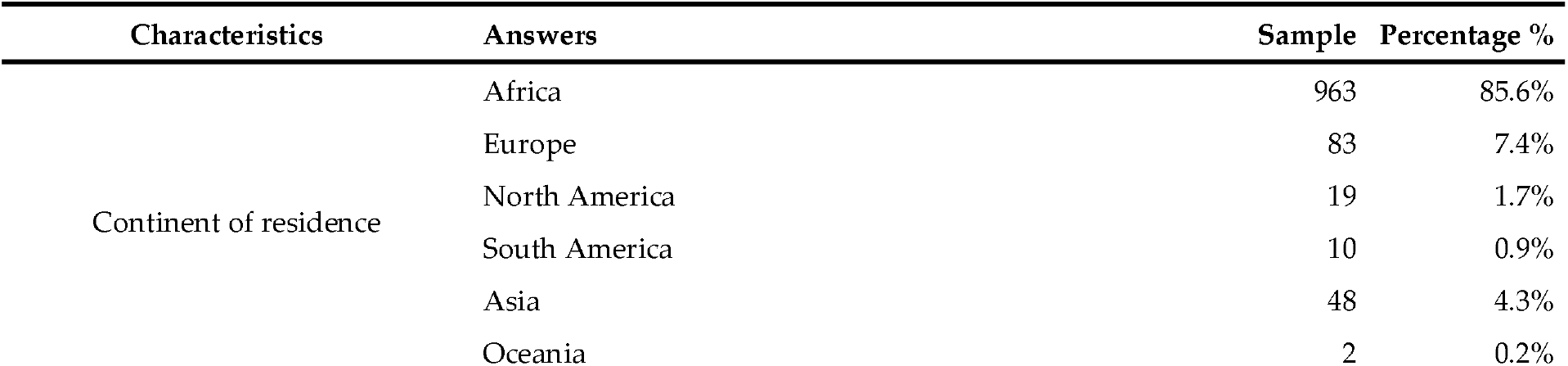
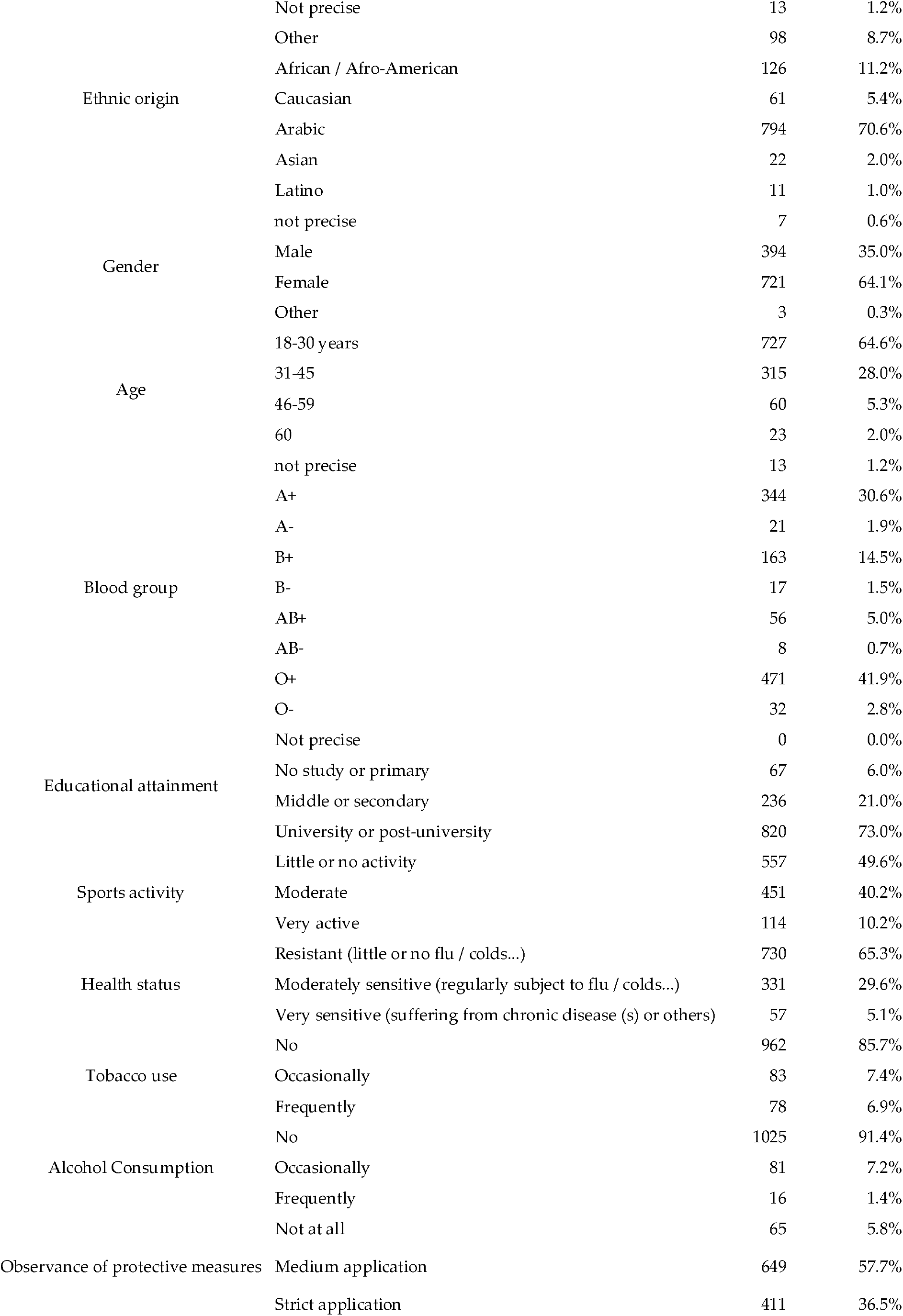
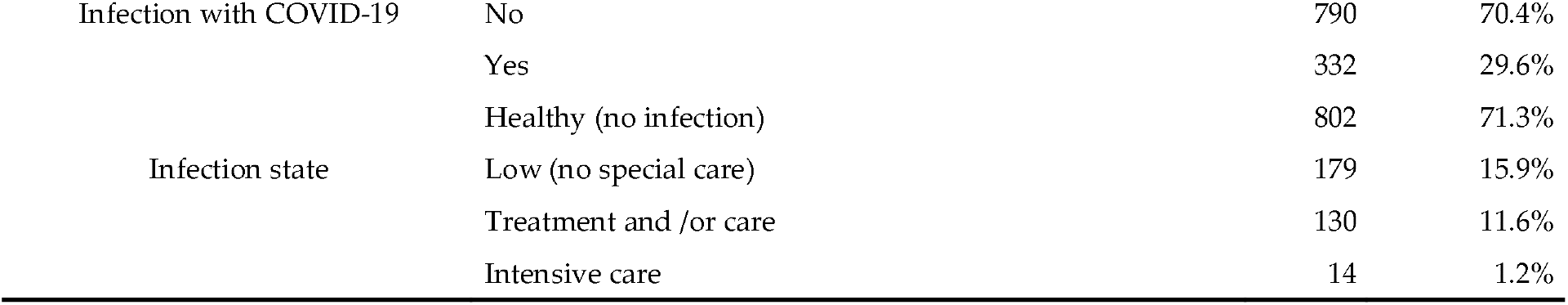
Characteristics of the sample study

### 3.2. Data analysis

Table 2 presents an overall image of analyses looking at the influence of considered predictors on the infection with COVID-19 and its severity. For each of the two, the table displays three models in separate columns. The first one looks at the individual relationship between the infection with COVID-19 and its severity with each predictor. The second column presents a “full model”, ascertaining the simultaneous influence of all predictors on the infection with COVID-19 and its severity. Finally, the third column presents a “prediction model”, resulted from the full one, and containing only predictors with a significant influence on the infection with COVID-19 and its severity, when considered simultaneously. The table clearly shows that age, sports and health status were significantly associated with the infection with COVID-19, whereas tobacco and protection were significantly associated with its severity.

**Table 2.** Analysis of the influence of considered predictors on the infection with COVID-19 and its severity. The table describes the influence, separately (column labeled “Univariate”), the simultaneously (column labeled “Full model”) and a final model resulting from selection including only predictors with a statistically significant simultaneous influence (column labeled “Selected model”). **Bold** values indicate a statistically significant impact (p≤0.05), and those in *Italic* a marginally significant impact (0.05<p≤0.1).

| Influence<br>(Model) | Dependent variable |  |  |  |  |  |
| --- | --- | --- | --- | --- | --- | --- |
|  | Infection |  |  | Severity |  |  |
|  | Univariate | Full model | Selected model | Univariate | Full model | Selected model |
| Continent | 0.3949 | *0.0891* |  | 0.2074 | *0.0888* |  |
| Ethnicity | 0.3217 | 0.1553 |  | 0.9307 | 0.6429 |  |
| Gender | 0.1654 | 0.7951 |  | 0.1088 | 0.7922 |  |
| Age | **0.0027** | **0.0185** | **0.0019** | 0.4958 | 0.6296 |  |
| Blood | *0.0667* | 0.1869 |  | 0.4963 | 0.4590 |  |
| Education | 0.2661 | 0.5919 |  | 0.6485 | 0.6156 |  |
| Sports | **0.0009** | **0.0010** | **0.0005** | 0.1478 | 0.2808 |  |
| Health | **0.0032** | *0.0919* | **0.0118** | *0.0723* | 0.1300 |  |
| Tobacco | *0.0934* | 0.6743 |  | **0.0164** | 0.3364 | **0.0370** |
| Alcohol | 0.1483 | 0.7615 |  | 0.1589 | 0.3068 |  |
| Protection | 0.4519 | 0.4083 |  | **0.0004** | **0.0042** | **0.0015** |

Having been affected or not by COVID-19 was analyzed via logistic regression whose results are presented in Table 3. The model was overall significant (p < 0.0001). The logistic regression demonstrated that age, sports activity and health status showed significant impacts on the infection of COVID-19.

**Table 3.**
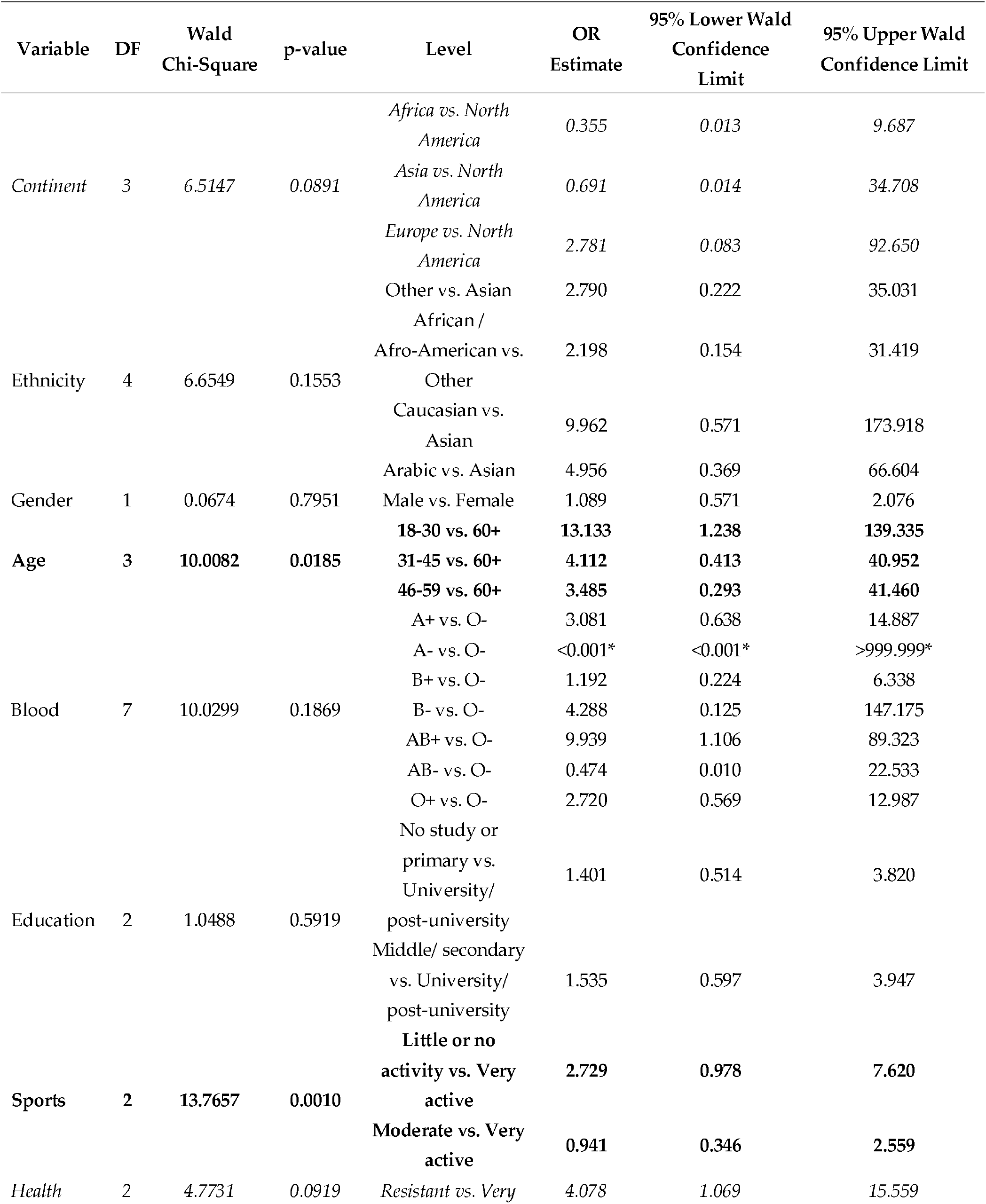
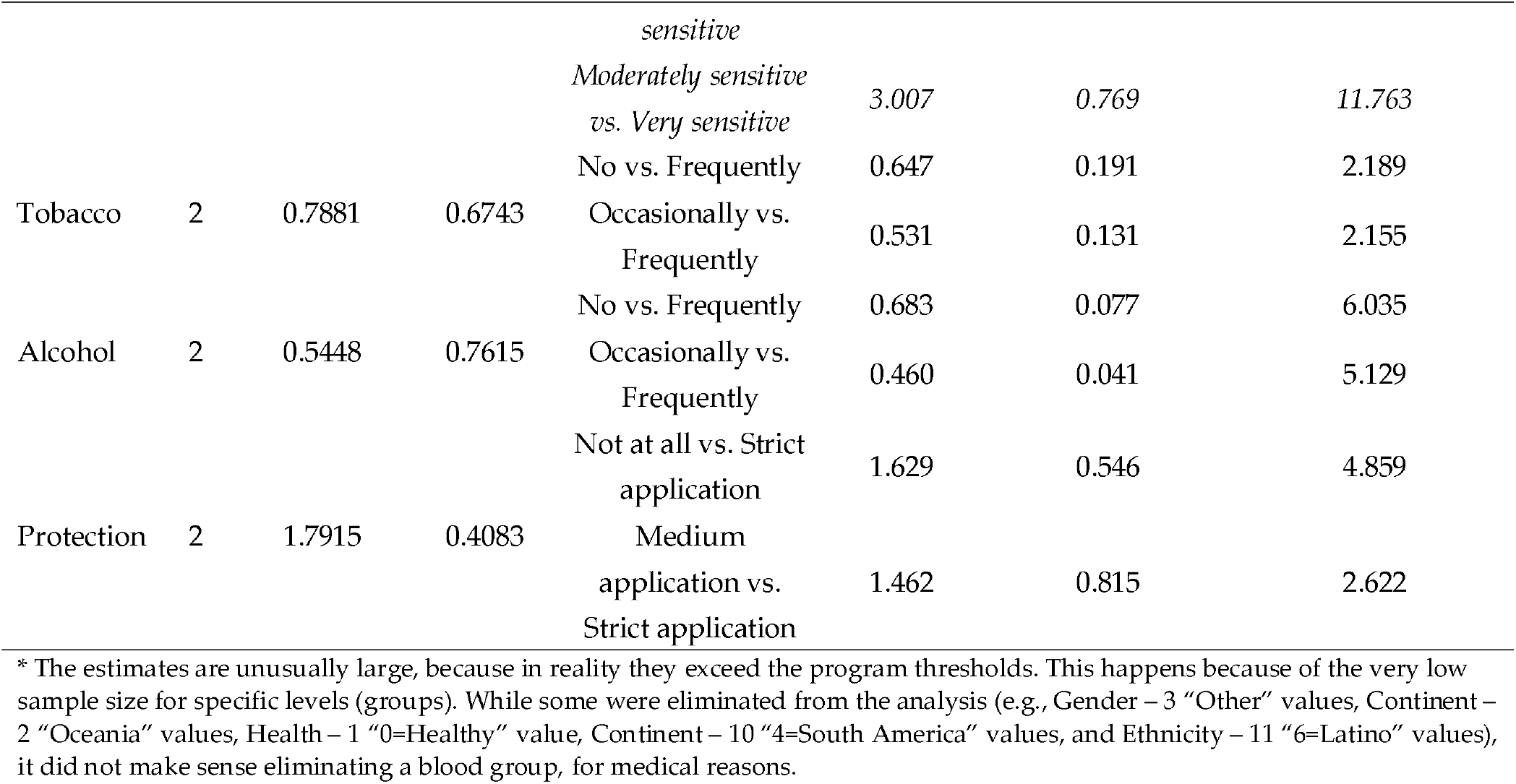
Results of the logistic regression showing the factors determining whether a subject has been affected by COVID-19 or not. **Bold** values indicate a statistically significant impact (p≤0.05), and those in *Italic* a marginally significant impact (0.05<p≤0.1). The model includes all analyzed factors.

Since not all factors in Table 3 were found to exert a statistically significant influence on having been affected or not by COVID-19, model selection was run in order to identify a model where all factors significantly affect the probability of having been affected or not by COVID-19. The model resulted by eliminating, in this order, the variables tobacco, gender, education, blood, and ethnicity. The resulting model, significant at p < 0.05, is displayed in Table 4.

**Table 4.**
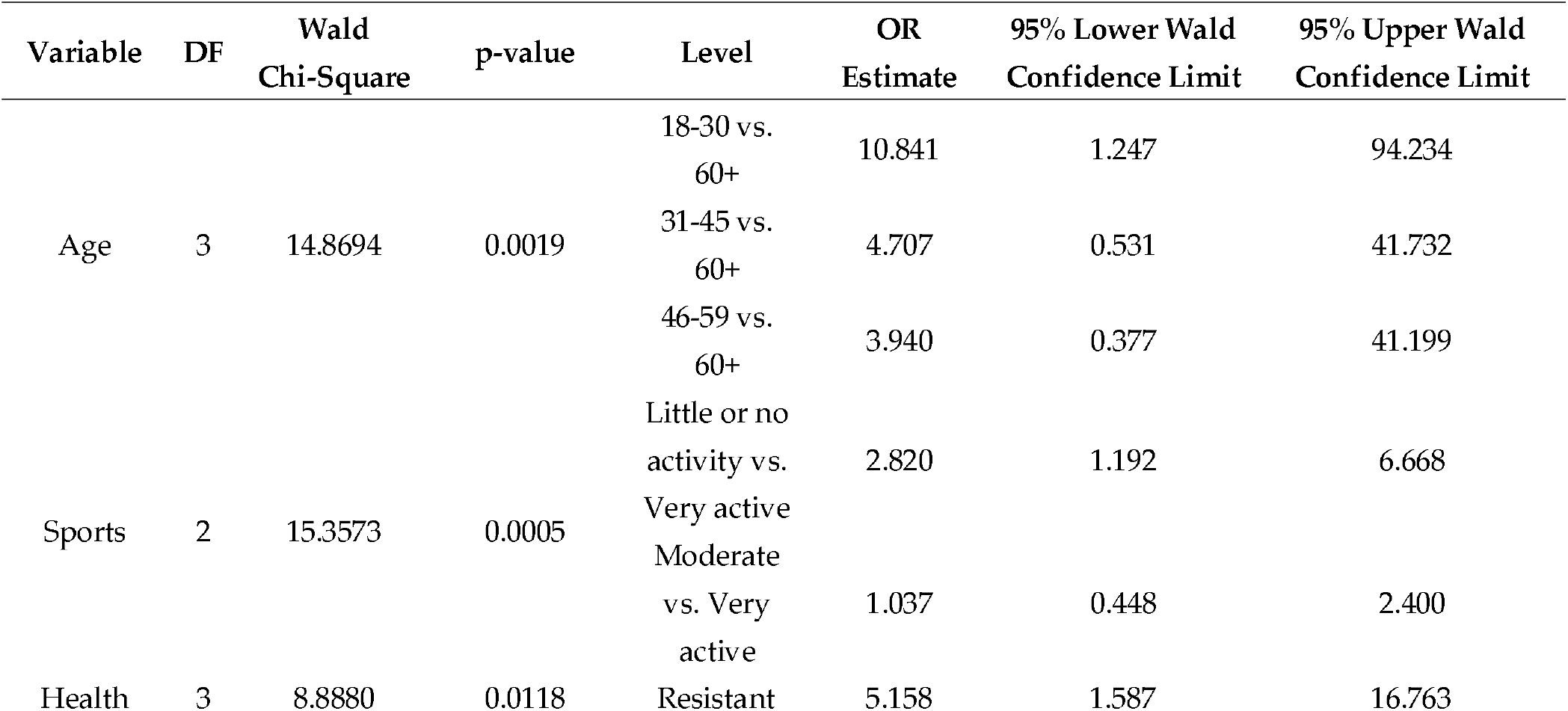
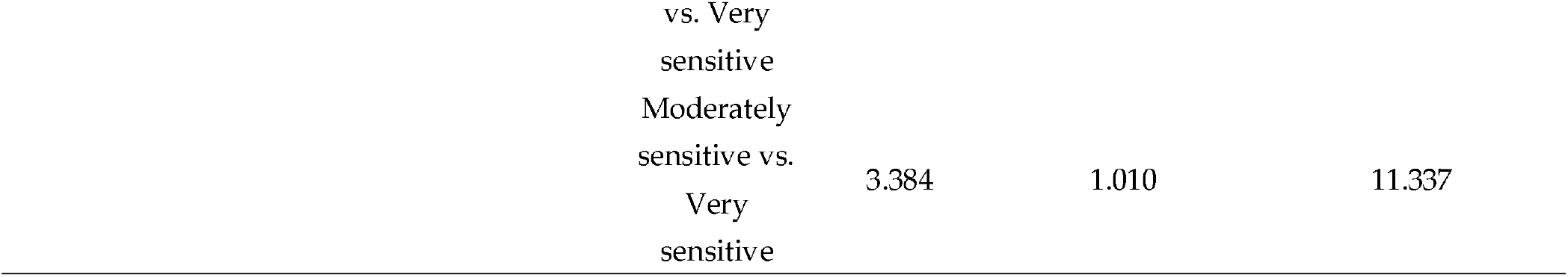
Results of the logistic regression showing the factors determining in a statistically significant way whether a subject has been affected by COVID-19 or not. All factors have a statistically significant impact (p≤0.05). Some levels were eliminated from the analysis (e.g., Gender – 3 “Other” values, Continent – 2 “Oceania” values, Health – 1 “0=Healthy” value, Continent – 10 “4=South America” values, and Ethnicity – 11 “6=Latino” values), but it did not make sense eliminating a blood group, for medical reasons.

The dependence of the severity of infection on potential risk factors investigated was analyzed using the logistic regression. The results are presented in Table 5. The model was overall significant (p = 0.0201). Here, the respect of protective measures and smoking were significantly associated with the severity of infection.

**Table 5.**
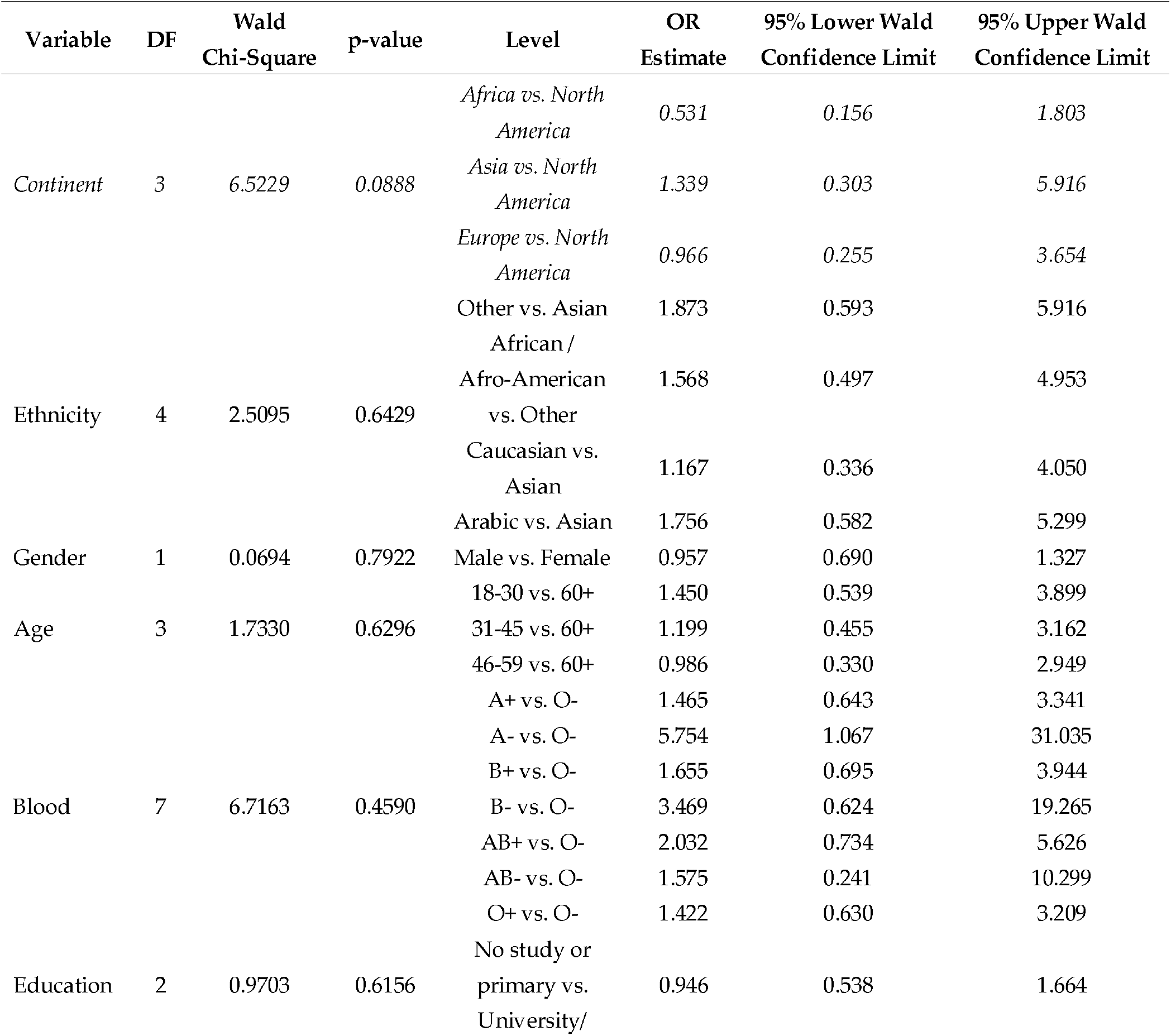
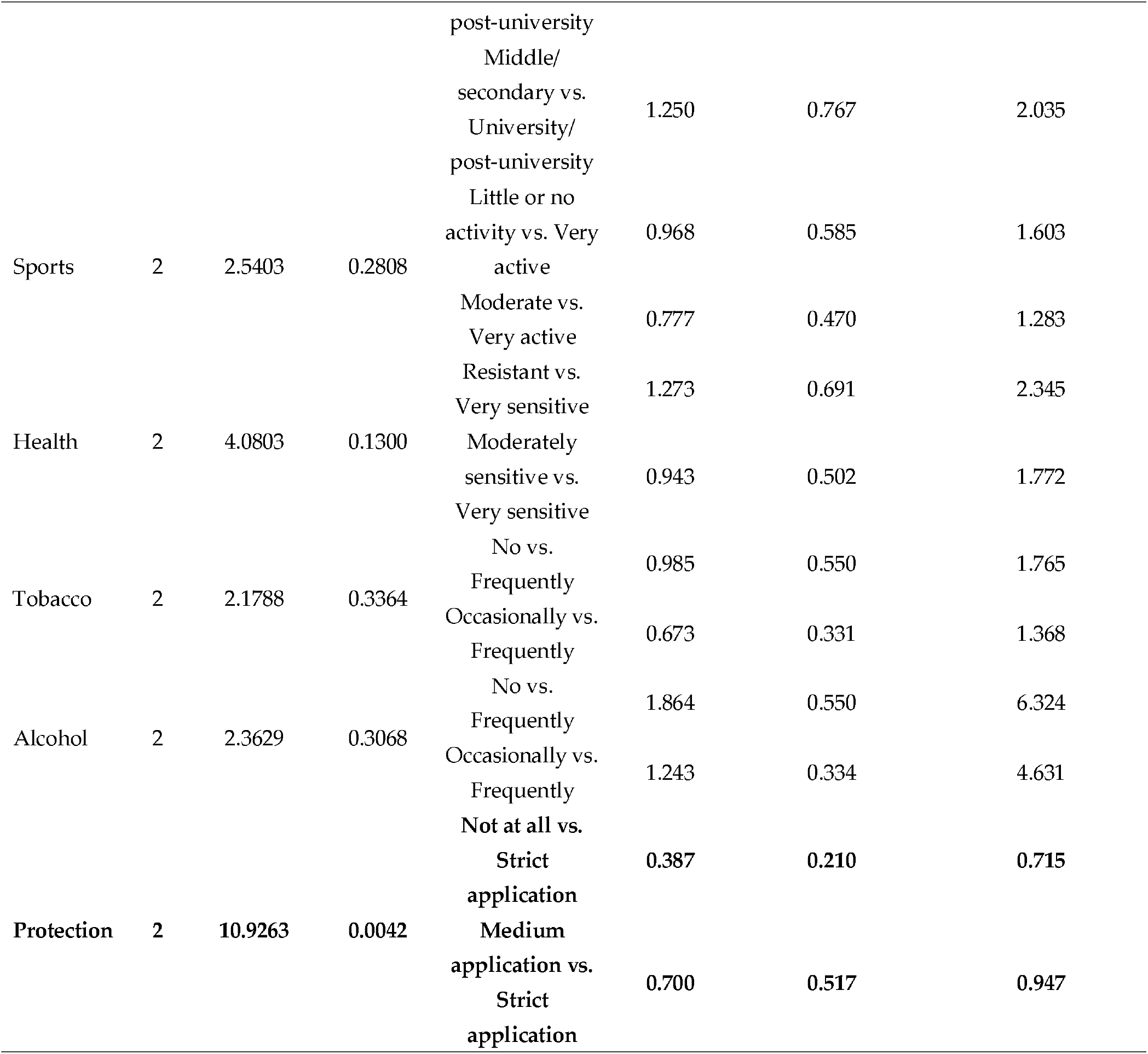
Results of the logistic regression showing the factors determining whether a subject has been severely affected by COVID-19 or not. Variables with names written in **Bold** have a statistically significant impact, and those in *Italic* a marginally significant impact (0.05<p≤0.1). The model includes all analyzed factors. Some levels were eliminated from the analysis (e.g., Gender – 3 “Other” values, Continent – 2 “Oceania” values, Health – 1 “0=Healthy” value, Continent – 10 “4=South America” values, and Ethnicity – 11 “6=Latino” values), but it did not make sense eliminating a blood group, for medical reasons.

Since not all factors in Table 5 were found to exert a statistically significant influence on the severity of COVID-19 infection, model selection was run in order to identify a model where all factors had a significant influence. The model resulted by eliminating, in this order, the variables gender, education, ethnicity, tobacco, blood, sports, continent, and age. The resulting model, significant at p < 0.05, is displayed in Table 6.

**Table 6.**
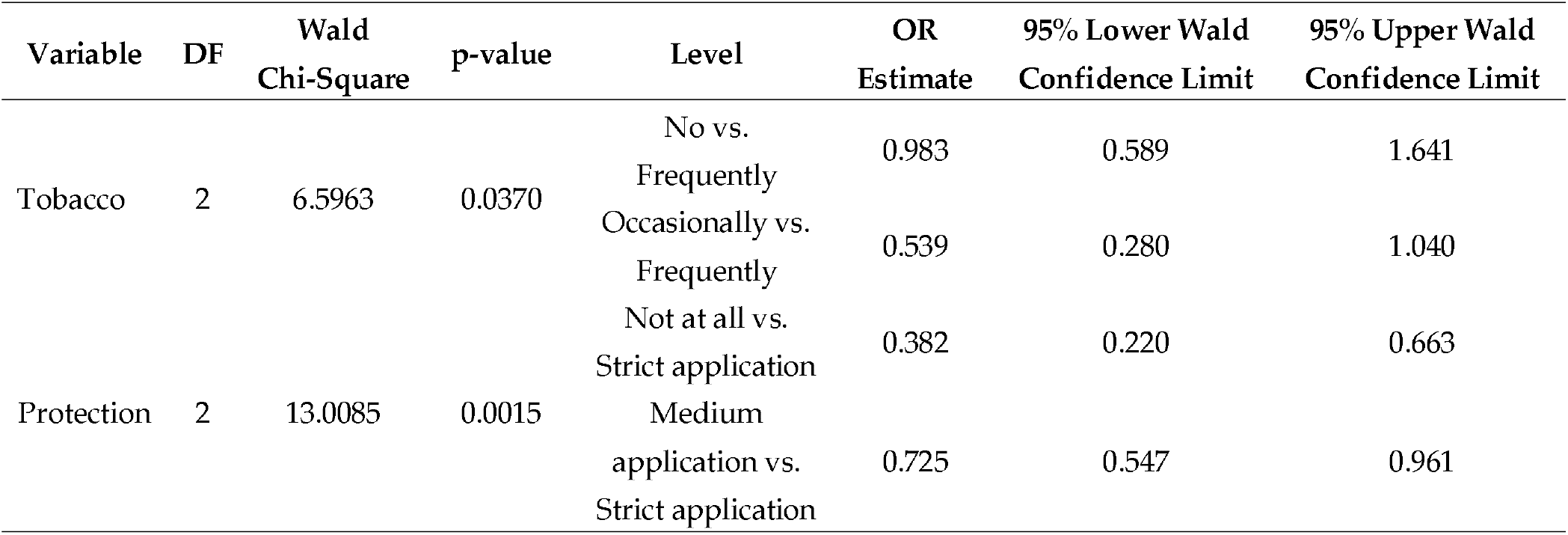
Results of the logistic regression showing the factors determining in a statistically significant whether a subject has been severely affected by COVID-19 or not. All factors have a statistically significant impact (p≤0.05). Some levels were eliminated from the analysis (e.g., Gender – 3 “Other” values, Continent – 2 “Oceania” values, Health – 1 “0=Healthy” value, Continent – 10 “4=South America” values, and Ethnicity – 11 “6=Latino” values), but it did not make sense eliminating a blood group, for medical reasons.

## 4. Discussion

### 4.1 Significance of the results and health recommendations

In this cross-sectional study, we evaluated association between innate and lifestyle-related factors on the rate and severity of COVID-19. The main findings are as follows: 1) age, physical activity, and health status were found to have a significant influence on the infection of COVID-19; 2) observance of preventive measures and tobacco consumption had a significant influence on the severity of the infection of COVID-19.

We confirmed the significant influence of the following factors on the possibility of infection: age, sports activity and to a lesser degree health status. We could thus interpret the factors on the rate of infection is by the fact that the older the person is, the more the individual will be subject to the infection, and this is confirmed with the positive wideness of the estimated parameter of the logistic regression, thus agreeing with previous studies [93,95]. With regard to physical activity, our results showed that the more active the individual, the less likely he is to be infected with the virus, this was also confirmed by the positive wideness of the estimated parameter of the logistic regression and agreeing with the previous studies [71,73]. While for the health status, the estimated parameter indicates that the more the individual’s sensitivity increases, the more the individual will be subject to infection, only confirming the results of previous studies concerning immunity [102]. Our results indicate that the compliance with preventive measures is essential. It seemed obvious to remind each time to observe the protective measures (wearing the mask, quarantine, remote working, hygiene, vaccination, etc.). The problem is that the average population thinks that after being vaccinated it is no longer mandatory to apply preventive measures [124]. It is true that previous studies have shown that vaccination of health care workers was associated with a substantial reduction in infections [125] or that one dose of vaccine reduces the potential for transmission by 61.0% [126], but the contamination possibility still exists. We encourage media and public health organizations to clarify this information [127].

In the same vein as for the probability of infection, it would seem for the severity that the more an individual smokes, the more severe his infection will be and will in most cases lead to hospitalization thus confirming the logic of previous work [56,57].With regards to protective measures, the logistic regression showed that the more the individual was strict in his application of the preventive measures promoted by the WHO and CDC [118], the less likely he will have a severe infection if he was infected. A previous online questionnaire observational study demonstrated that the use of protective measures was not associated with symptom severity of COVID-19 [128]. Another South Korean study showed that preventive measures had a negative influence on cardio metabolic profiles in subjects with metabolic impairments [129]. To our knowledge, there has not been any work that has demonstrated the positive effect of preventive measures on COVID-19. In contrast, a study in Pakistan showed that the application of protective measures dramatically reduces the risk of other respiratory diseases [130] Recently a Japanese study focused only on wearing a mask as a measure and they found that for a sample of 820 individuals, mask users were infected at a rate of 0.4 times that of those who did not wear masks [131] demonstrating the importance of this measure. Our findings provide novel insights into the role of personal protective measures on the spread of infection and symptom severity. Further study is warranted to confirm our findings.

Regarding age, since the beginning this pandemic, COVID-19 has always been associated with old individuals [132]. A higher need for intensive care increase by age among individuals older than 45 years [133]. Our findings only confirm the importance of this factor by going in the same direction as previous reviews and meta-analysis studies [134,135].

Concerning alcohol, the situation is a little paradoxical. Our findings showed that the alcohol consumption has no significant influence on the probability of infection and its severity. However, Abbasi-Oshaghi et al. [136] found the opposite, because long-term drinking is closely related to zinc and vitamin deficiency. Hikida et al. [137] also found out that alcohol probably increases the risk of exposure to and morbidity from COVID-19. In addition, according to the same study, alcohol consumption is associated with prosecution and having regrets, which can increase the severity of COVID-19. González-Reimers et al. [138] discussed the fact that it has already been proven that various chronic disorders may increase the risk of pneumonia (and by extrapolation COVID-19) severity. Precisely for the severity, a Chinese study performed on more than 1,500 patients claimed that the severity of infection was greater in patients who consume alcohol regularly or with a history of consumption [139]. At the same time, a recent review article also confirmed that alcohol consumption adds vulnerability to patients and leads to an increased severity of COVID-19 and other long-term health problems [140]. Nevertheless, others remain a little less categorical leaving doubt by pointing out the influence of several factors at the same time as well as the need for further studies [141,142].

While for tobacco, the results of our work are quite logical because it was more associated with the severity of infection compared to the infection. Indeed, non-smokers are obviously not immune to the virus; however, smoking patients are vulnerable to severe COVID-19 [143]. This perfectly confirms the findings of previous studies [144,145].

In general, the measures must be subject to a special focus especially for sensitive sectors. Citing as an example the education sector, where the application of measures was investigated in a few articles [146,147] but the most striking example remains, as stated previously, the health sector [148,149]. We think that it is important for each individual to educate about his own health on a regular basis. This issue remains debatable, as in many countries the relationship between educational disparities and socioeconomic status is discussed a lot without reaching a final answer [150,151]. Variability in immune system components (innate) is a major contributor to the heterogeneous disease courses noticed for Sars-CoV-2 [152]. The immune response differs from a person to another and should not be relied on in any way provided the still unknown character of several mechanisms [153]. A continuous effort to understand all mechanisms, especially those concerning host-virus interactions, is essential to overcome the pandemic [154]. In our study, a health state was associated with a reduced probability of contracting the virus, which can demonstrate the importance of innate immunity, but in no case should we be totally based on this. Nevertheless, we cannot limit human health solely to these factors, and the effectiveness of vaccine plays the most important role. Further studies are needed to confirm the relationship between vaccines, pollution, and their consequences [155].

A point not to be overlooked concerns the sources of information (or disinformation). A recent study on Jordanian and Iraqi participants revealed that more than half of the population of the two countries use social media as the most common COVID-19 information source, sharing a lot of false information which may lead to gaps in the actions to be adopted [156]. A cross-sectional study conducted in Saudi Arabia focused on testing the knowledge of residents with questions about the measures to be taken, and more than 90.0% of the population was above average by specifying in particular that people with a low level of education have failed [157]. Still in the same country, the observance of precautionary measures among nursing staff was studied, and results show that a high number (71.2%) were competent and less qualified personnel should nevertheless be trained in particular [158]. This would be applicable to all countries, and could prove useful.

Finally, our original research goal was to analyze the influence of two groups of factors and their influence on COVID-19 to reduce the spread of the virus. These findings also agree with the WHO reports who have constantly repeated the various protective measures to be taken for avoiding any possible infection and thus indirectly limiting the spread of virus [6].Additionally, this study is one of only few going against the idea that a part of the world population is automatically condemned to infection because of their ethnicity, place of residence etc. Our study is meant to be encouraging by showing that each person has almost entirely its destiny in its hands as long as a healthy lifestyle is observe and precautions taken; it is also about advising people to self-educate. This can have only positive effects, even in the long term.

### 4.2 Importance of the study

The originality of this study comes from the fact that it is the only international study providing more information on the African and Mediterranean areas, carried out in 4 different languages. The number of respondents is quite high and heterogeneous compared to almost all questionnaires presented in previous studies. Moreover, the vast majority of publications have focused on a particular sector [159], a particular region [160] or a well-targeted population [161]. In addition, previous studies dealt with a single characteristic or group of characteristics [162,163], while the present study includes 13 items based on a substantial literature review. These items addressed two groups of characteristics comparing the most significant ones. Everything was simplified as much as possible in order to reach all categories of the world population and collect more information applicable to the vast majority, so that we can encourage them to adopt the right actions to prevent the infection, and ultimately limit the spread of disease. Moreover, the ranked influence of COVID-19 infection drivers has never been evaluated before. Any additional findings are important, provided their possible contribution to the fight against this pandemic.

### 4.3 Methodological limitations

Our analysis was limited to a number of factors considered as relevant based on the literature review, and which could be ascertained using an online questionnaire. However, different studies also pointed out a number of other potential risk factors.

Objectively, it is an almost impossible challenge to know exactly the factors responsible for infection and transmission of COVID-19. Sources may be incomplete; apart from the factors discussed previously, even the meteorological ones were considered a potential explanation [164]. A study in Korea demonstrated that the environment plays a significant role in the spread of COVID-19, but like any factor, it may have also been impacted by various additional features [165]. Hence, further studies are needed to protect people from COVID-19 transmission, specifically on infection dynamics and the mode of transmission, e.g., cluster spaces, closed spaces, and indoor environments [166].

At the individual level, everyone must take the maximum possible precautions. It should also be remembered that no less than 10 reasons supporting airborne transmission were phrased recently by Greenhalgh et al. [167]. The long-term health consequences of COVID-19 remain unclear and continue to be studied [168]. Therefore, it is preferable to avoid any form of infection, even mild. Another factor that we do not necessarily think about and which may be important is the wastewater treatment and disinfection strategies with chemical products; all demonstrate that we must take into account an incalculable number of factors [169].

We can also mention certain biases concerning the sample. The majority of respondents were young intellectuals from the Arab (and Muslim) world; consequently, tobacco and alcohol remain taboo subjects, so it is possible that some respondents did not tell the truth about these characteristics, and our results could be biased. Integrating vaccination as a protective measure rather than as a factor in its own right when carrying out this sampling was not a limiting factor; time has showed that this is the case now.

### 4.4 Perspectives for future research

Undoubtedly, this study could not explicitly pretend to yield results able to stop this pandemic. In general, making predictions on this virus, which remains unknown, is impossible. Therefore, any additional information may prove useful. We consider that each study on this topic contributes in its own way to fighting indirectly against the pandemic crisis we are currently experiencing. Previously, we have enumerated a certain number of limitations in this work, including the number of answers received to the questionnaire. While this is acceptable, more answers could help obtaining more representative results, and increasing their statistical significance. Future studies should seek to use a larger and even more diverse sample, and include additional recruitment strategies. For example, future studies should include in their sample information provided by parents of children under 18 years old, as they are also a group of high concern given the target specificity of the new virus variants.

## 5. Conclusions

In this study, we found that health status, age and physical activity significantly influence the infection (p < 0.05). The non-respect of preventive measures and smoking tobacco significantly aggravate its severity (p < 0.05). It would also appear that no matter where an individual is, what his/her age is, and everything that characterizes the person from birth, are not ultimately the main characteristics to be taken into consideration concerning the COVID-19 infection (and severity). Despite the limitations, based on our results this is the first study that demonstrated that it would be much more the lifestyle of the individual that decides a potential infection. Therefore, we strongly encourage the world population to adopt a healthy lifestyle: no alcohol, no tobacco, or at least reducing their consumption; eat healthily, sleep well and practice regularly a physical activity, in addition to a massive vaccination, specifically for medical staff and people at risk. Finally, the capital points not to be neglected will inevitably remain the fact of avoiding as much as possible any gathering (apply social distancing, in particular by using remote working to the maximum extent possible) and observe the protective measures promoted by the WHO. Further studies remain necessary to help fighting against the constantly evolving epidemiological situation. In the end, we hope that this positive message that we deliver (supported by results) can positively influence the behavior of people while waiting for the health crisis to subside and for the world to return to normality.

## Data Availability

All data produced in the present study are available upon reasonable request to the authors

## Author Contributions

Conceptualization, H.A.A., M.A. and A-I.P; methodology, H.A.A., M.A. and A-I.P.; validation, C.M.L., L.E. and N.O.; formal analysis, H.A.A, A-I.P., L.E. and N.O.; investigation, H.A.A. and M.A.; resources, A.S., M.S.A.K.; data curation, M.A and A-I.P., writing—original draft preparation, H.A.A, A-I.P., J.Y. and L.E.; writing—review and editing, H.A.A, A-I.P., A.S., J.Y. and N.O.; visualization, C.M.L., L.E and N.O. supervision, A-I.P, L.E. and N.O.; project administration, M.S.E.K.; funding acquisition, H.A.A., A.S., and M.S.E.K. All authors have read and agreed to the published version of the manuscript.

## Institutional Review Board Statement

Not applicable.

## Informed Consent Statement

Informed consent was obtained from all subjects involved in the study and all procedures were approved by the CRSTRA.

## Data Availability Statement

The data generated and/or analyzed in the current study is available from the corresponding author on reasonable request.

## Acknowledgments

Considerable appreciation is addressed to the DGRSDT (Directorate-General of Scientific Research and Technological Development of Algeria) and the MESRS (Ministry of Higher Education and Scientific Research of Algeria) for their valuable support. The authors thank, as well, all of the respondents in this study for their voluntary participation and for providing such essential information. Many thanks are also addressed to anyone who helped us by sharing the questionnaire.

